# Pneumococcal exposure routes for infants, a nested cross-sectional survey in Nha Trang, Vietnam

**DOI:** 10.1101/2021.07.04.21259950

**Authors:** George Qian, Michiko Toizumi, Sam Clifford, Lien Thuy Le, Tasos Papastylianou, Billy Quilty, Chihiro Iwasaki, Noriko Kitamura, Mizuki Takegata, Trang Minh Nguyen, Hien Anh Thi Nguyen, Duc Anh Dang, Albert Jan van Hoek, Lay Myint Yoshida, Stefan Flasche

## Abstract

**Background:** Infants are at highest risk of pneumococcal disease. Their added protection through herd effects is a key part in the considerations on optimal pneumococcal vaccination strategies. Yet, little is currently known about the main transmission pathways to this vulnerable age group.

**Methods and findings:** We conducted a nested cross-sectional contact and nasopharyngeal swabbing survey in randomly selected infants across all 27 communes of Nha Trang, Vietnam. Bayesian logistic regression models were used to estimate age specific carriage prevalence in the population, a proxy for the probability that a contact of a given age could lead to pneumococcal exposure for the infant. We used another Bayesian logistic regression model to estimate the correlation between infant carriage and the probability that at least one of their reported contacts carried pneumococci, controlling for age and locality. In total 1583 infants between 4 and 13 months old participated, with 7428 contacts reported. Few infants (5%) attended day care and carriage prevalence was 22%. Most infants (61%) had less than a 25% probability to have had close contact with a pneumococcal carrier on the surveyed day. Pneumococcal infection risk and contact behavior were highly correlated: if adjusted for age and locality the odds of an infant’s carriage increased by 22% (95%CI:15-29) per 10 percentage points increase in the probability to have had close contact with at least one pneumococcal carrier. Two to six year old children contributed 51% (95%CI: 39-63) to the total pneumococcal exposure risks to infants in this setting.

**Conclusions:** Cross-sectional contact and infection studies can help identify pneumococcal transmission routes. In Nha Trang, preschool age children are the largest reservoir for pneumococcal transmission to infants.

## Introduction

Mathematical modelling is a key part of the evidence synthesis process that informs public health policy for infectious diseases and their mitigation, not least evident in its role in the SARS-CoV-2 pandemic [1,2]. An essential part of such infectious disease models is their adequate reflection of transmission in the community studied. For respiratory pathogens, social contacts and their age structure has been used predominantly as the key source of heterogeneity in infectious disease spread and hence the accurate measurement of social contacts as a proxy for potential transmission events has been fundamental for the validity of such models [3,4].

While social contacts are a seemingly obvious proxy for transmission risk for pathogens transmitted via droplets or aerosol from the respiratory tract, there is limited direct evidence that the common definition of a transmission relevant contact, i.e. a two way conversation or skin-to skin contact, is indeed a major risk factor for infection. Nested contact and infection studies in Fiji and Uganda have shown that the frequency of physical (skin-to-skin) household contacts of longer duration (>1 hour) were associated with higher rates of pneumococcal carriage [5,6] and a prospective cohort study in the UK showed that adults hospitalised with community acquired pneumonia had increased odds for close child contact within the 4 weeks prior to their illness [7]. Furthermore, during the SARS-CoV-2 pandemic changes in contact behaviour as measured by social contact surveys have enabled real time estimation of the impact of non-pharmaceutical interventions on pandemic transmission intensity [8,9].

Infants are typically at high risk for severe respiratory disease and are targeted for either direct or indirect vaccine protection. Their exposure to disease through their social contacts can also be an important guide for selecting different dosing schedules for these vaccines, including pneumococcal conjugate vaccines [10]. Yet, there is little evidence on what constitutes a transmission relevant contact for infants or the frequency and distribution of such contacts. Most contact studies include few infants [3,9] or are conducted in specific environments that capture only parts of their contacts [11]. The most detailed study to date, which studied the contact patterns of 115 infants in the UK, found that almost two thirds of infant contacts were not with household members, highlighting the potential importance of non-household transmission routes for infant infection [12].

We report the results of a large infant contact study nested into a cross-sectional pneumococcal carriage survey in Nha Trang, Vietnam. We investigate the correlation of infant exposure risk, approximated by reported social contacts and the prevalence of infection in such, with their risk of pneumococcal infection and the spatial and demographic structure of infant contacts and exposure in this setting.

## Methods

### Study population

The study was conducted in Nha Trang, a coastal city in south-central Vietnam with a total population of just over 426,958 and under-five population of 28,495 in the 2018 census (personal communication with Khanh Hoa Health Service). Nha Trang city consists of 27 communes of similar population size. Each commune has a commune health centre, providing a range of basic health services. Similarly, educational services including nurseries and schools are largely provided on a commune basis.

In October 2016 a cluster randomised controlled trial was initiated to evaluate alternative pneumococcal conjugate vaccine (PCV) dosing schedules [13]. In the 24 communes that were allocated to receive PCV, routine infant vaccination was initiated according to the allocated schedule and a catch-up campaign including children less than three years old was conducted. The primary endpoint of the trial relies on pneumococcal carriage and, as such, is monitored in each commune through annual cross-sectional nasopharyngeal carriage surveys in 60 infants (4 months - 11 months), 60 toddlers (14 months - 23 months) and their respective main carers.

### Study design

In October 2018, two years after the start of the trial, an infant contact survey was nested into the cross-sectional carriage study. All infants enrolled for the carriage study were also eligible for enrollment into the contact survey.

For each commune health centre, a staff member was identified and trained to conduct the contact survey in their local commune. During an initial home visit, written informed consent was obtained from parents or guardians before any collection of data or specimen. Subsequently a background questionnaire was filled in together with the caretaker on behalf of the infant, collecting information including sex, age, household composition and the infant’s mobility (See SIFile 1 for the full questionaire). The carer was asked to monitor their infant’s contacts the following day and another home visit was scheduled for the day after. For that visit the interviewer would aid the carer’s memory by discussing their day and noting initials of the infant’s contacts. Subsequently, information on those contacts’ age, gender, duration and location was filled in (See SIFile 2 for the full questionnaire). A contact was defined as skin to skin contact with the infant, because the common conversational contact definition would be inappropriate to apply to infants and because physical contacts, as a proxy for close contacts, have been shown to better correlate with pneumococcal carriage risk [5,6].

During the first home visit for the contact study, consent to participate in the nasopharyngeal carriage study was sought from the carers. Participants subsequently were given an appointment with the commune health centre during which a nasopharyngeal swab from the infant was obtained by a study nurse. Sample collection, handling and storage were performed according to World Health Organization guidelines [14]. At Pasteur Institute of Nha Trang real-time qPCR was performed detecting the autolysin-encoding gene (lytA) of Streptococcus pneumoniae. Microbiological culture was subsequently attempted for lytA positive (cycle threshold (Ct) value < 35) or equivocal (Ct value 35-40) samples.Pneumococcal carriage was defined as samples that were both lytA positive/equivocal and culture positive, and subsequently confirmed by microarray [15].

### Statistical analyses

We used negative binomial regression to estimate crude rate ratios of the mean number of contacts of participants in different population subgroups. We also calculated rate ratios adjusted for age (categorical: 4-7 months and 8-11 months of age) and for clustering of participants into communes.

We used logistic regression to estimate the crude ratio of odds for a contact being made outside the commune of residence across population subgroups. In an adjusted analysis we included the age group (4-7 and 8-11 months of age) and day-care attendance as covariates, and accounted for clustering of participants into communes.

To explore whether an infant’s contact patterns are correlated with their probability to carry pneumococcus, we created the Potential Exposure Index (PEI). For each infant participating in the study, PEI estimates the probability that at least one of the reported contacts was carrying pneumococcus and hence that the infant was exposed to the pneumococcus on that day at least once. We define PEI as:

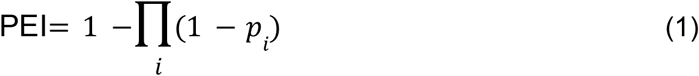

where *p*_*i*_ represents the probability that the *i*th contact of the respective study participant is a pneumococcal carrier. We assume that *p*_*i*_ is solely dependent on the contact’s age. To estimate age dependent probabilities for pneumococcal carriage in the study setting, we performed Bayesian Penalised B-Spline Regression on logit-transformed carriage probability, obtained from previously reported low-granularity age-stratified pneumococcal carriage across age groups from Nha Trang [16]. We compared our estimates against carriage prevalence observed in young children in the trial. The PEI estimate is based on the contact patterns recorded for a single day but were interpreted as a proxy for the general contact behaviour of the respective infant.

We used a hierarchical Bayesian logistic regression model (see SIFile 3) to estimate the correlation of PEI (covariate) and pneumococcal infection (outcome). Based on selection via a Bayesian Belief Network (see SIFigure S4 for the Directed Acyclic Graph), we included age in months as a covariate and the resident community as a random effect. The model was implemented in JAGS [17]. The uncertainty in the individual level estimates for PEI in this analysis was retained from the posterior estimates in the previous regression (namely, the cubic spline on age) by assuming a normal distribution for each year of age and drawing samples from this distribution when calculating the posterior samples of PEI. We chose to assume a normal distribution because for simplicity and since, by visual inspection, the distribution of carriage rates across all ages appeared to follow a Gaussian. Subsequently, this individual-level uncertainty in PEI is further propagated by assuming a beta distribution (again, by visual inspection) for these PEI estimates; this distribution then serves as an input to the logistic regression predicting carriage status. Potential correlations among the predictor variables were checked before model fitting using the Variance Inflation Factor (VIF) [18].

The total pneumococcal exposure to an infant on the day that contacts were reported was approximated by the sum of each reported contact’s age specific probability of pneumococcal carriage. The contribution of a specific age group to that total was calculated as the pneumococcal exposure from contacts of that age group divided by the total exposure. This can be generalised to calculate the contribution of specific age groups to the total pneumococcal exposure across all infants by including all contacts in the study.

The descriptive statistics were done in STATA version 14.0 [19]. The analyses on the association of contact patterns and pneumococcal carriage were conducted in R 4.0.2 [20] and RJags [21]. The code is available on: https://github.com/GeorgeYQian/Nha-Trang-Contact-Study-Data-and-Code

### Ethical approval

Ethical approval for the study was granted by respective ethics boards of National Institute of Hygiene and Epidemiology (4405/QD-BYT) and the London School of Hygiene & Tropical Medicine (15881).

## Results

### Characteristics of study participants and their contacts

Overall, 1583 infants aged between 4 and 13 months (median: 9, interquartile range 7-11 months) were enrolled in both the contact survey and the carriage survey (Table 1). All children were not older than 11 months at the time of enrollment but 122 children were 12 or 13 months at the time of data collection because of delays between enrollment and data collection. An additional 11 infants were enrolled in the carriage survey but did not consent inclusion for the contact survey; these were not included in any of the following analyses. 58% (917) infants had siblings living in the same household and the average household size was 4.9 (standard deviation 1.0). The majority of infants were able to sit on their own (84%), many could crawl (55%) and some walk (12%). Most families had a motorbike (98%) but other means of transport were rarely used. 50% of infants did not leave their commune of residence during the preceding week. Few infants (5%) attended day care. Pneumococcal carriage prevalence was 22% and increased from 14% at 4-6 months to 27% at 11-13 months.

**Table 1:** Characteristics and mean number of contacts reported for all infants enrolled into both the carriage and the contact study.

| Characteristic | Number | Mean (SD) | Ratio | Adjusted ratio* |
| --- | --- | --- | --- | --- |
| Total | 1583 | 4.7 (1.8) |  |  |
| Demographics |  |  |  |  |
| Sex |  |  |  |  |
| Male | 871 | 4.7 (1.8) | reference | reference |
| Female | 712 | 4.6 (1.8) | 0.98 (0.94-1.03) | 0.98 (0.94-1.01) |
| Age (months) |  |  |  |  |
| ≤7 months | 616 | 4.7 (1.8) | reference |  |
| 8-13 months | 967 | 4.7 (1.8) | 1.01 (0.97-1.06) |  |
| Family |  |  |  |  |
| Siblings in the household |  |  |  |  |
| No sibling | 666 | 4.6 (1.9) | reference | reference |
| One or more siblings | 917 | 4.7 (1.7) | 1.03 (0.98-1.08) | 1.03 (0.99-1.06) |
| Number of people in the household |  |  |  |  |
| 2-4 | 611 | 4.1 (1.7) | reference | reference |
| >4 | 972 | 5.1 (1.7) | 1.25 (1.19-1.31) | 1.25 (1.19-1.30) |
| Caretaker currently in paid employment |  |  |  |  |
| No | 1056 | 4.7 (1.9) | reference | reference |
| Yes | 527 | 4.7 (1.6) | 1.01 (0.96-1.06) | 1.01 (0.96-1.06) |
| Highest level of education in the household |  |  |  |  |
| Up to secondary school | 268 | 4.8 (1.8) | reference | reference |
| Degree | 1315 | 4.7 (1.8) | 0.98 (0.93-1.04) | 0.98 (0.92-1.05) |
| Infant's activity |  |  |  |  |
| Sit |  |  |  |  |
| Yes | 1334 | 4.7 (1.8) | 1.04 (0.98-1.11) | 1.05 (0.99-1.11) |
| No | 249 | 4.5 (1.7) | reference | reference |
| Crawl |  |  |  |  |
| Yes | 873 | 4.7 (1.8) | 1.01 (0.96-1.05) | 1.00 (0.95-1.05) |
| No | 710 | 4.7 (1.8) | reference | reference |
| Walk |  |  |  |  |
| Yes | 186 | 4.7 (2) | 0.99 (0.93-1.07) | 1.10 (0.98-1.24) |
| No | 1397 | 4.7 (1.8) | reference | reference |
| Mobility |  |  |  |  |
| Bicycle |  |  |  |  |
| Yes | 31 | 4.5 (1.4) | 0.95 (0.80-1.12) | 0.95 (0.86-1.04) |
| No | 1552 | 4.7 (1.8) | reference | reference |
| Motorbike |  |  |  |  |
| Yes | 1549 | 4.7 (1.8) | 1.15 (0.97-1.36) | 1.15 (1.06-1.25) |
| No | 34 | 4.1 (1.2) | reference | reference |
| Car |  |  |  |  |
| Yes | 99 | 4.9 (2.1) | 1.04 (0.95-1.14) | 1.04 (0.97-1.13) |
| No | 1484 | 4.7 (1.8) | reference | reference |
| Walk |  |  |  |  |
| Yes | 108 | 5.1 (2.0) | 1.1 (1.01-1.20) | 1.1 (0.98-1.24) |
| No | 1475 | 4.7 (1.8) | reference | reference |
| Public transportation |  |  |  |  |
| Yes | 40 | 4.8 (1.8) | 1.03 (0.89-1.19) | 1.03 (0.90-1.18) |
| No | 1543 | 4.7 (1.8) | reference | reference |
| Number of times caretaker left the commune in the last 7 days |  |  |  |  |
| 0-2 | 872 | 4.5 (1.7) | reference | reference |
| 3 or more | 711 | 4.9 (1.9) | 1.07 (1.03-1.12) | 1.07 (1.03-1.12) |
| Number of times infant left the commune in the last 7 days |  |  |  |  |
| 0 | 788 | 4.6 (1.7) | reference | reference |
| 1 or more | 795 | 4.8 (1.8) | 1.06 (1.01-1.11) | 1.06 (1.02-1.10) |
| Day-care |  |  |  |  |
| Day-care attendance |  |  |  |  |
| Yes | 86 | 5 (1.5) | 1.07 (0.97-1.18) | 1.07 (0.999-1.14) |
| No | 1497 | 4.7 (1.8) | reference | reference |
| Pneumococcus carriage (n=1582) |  |  |  |  |
| Pneumococcus |  |  |  |  |
| Yes | 353 | 4.8 (1.8) | 1.03 (0.97-1.09) | 1.03 (0.98-1.08) |
| No | 1229 | 4.7 (1.8) | reference | reference |

\*Incidence rate ratios adjusted by age group, considering clustering in each commune

A total of 7428 contacts aged between 0 and 100 years (median: 33 years) were reported (Figure 1). Contacts mostly occurred at home (89%) and with family members (76%). They were mostly with other children, parents and grandparents and lasted for more than 1 hour (55%). Few contacts (5%) were reported to occur outside the commune of residence or with other infants (0.4%) (Table 2).

**Table 2:**
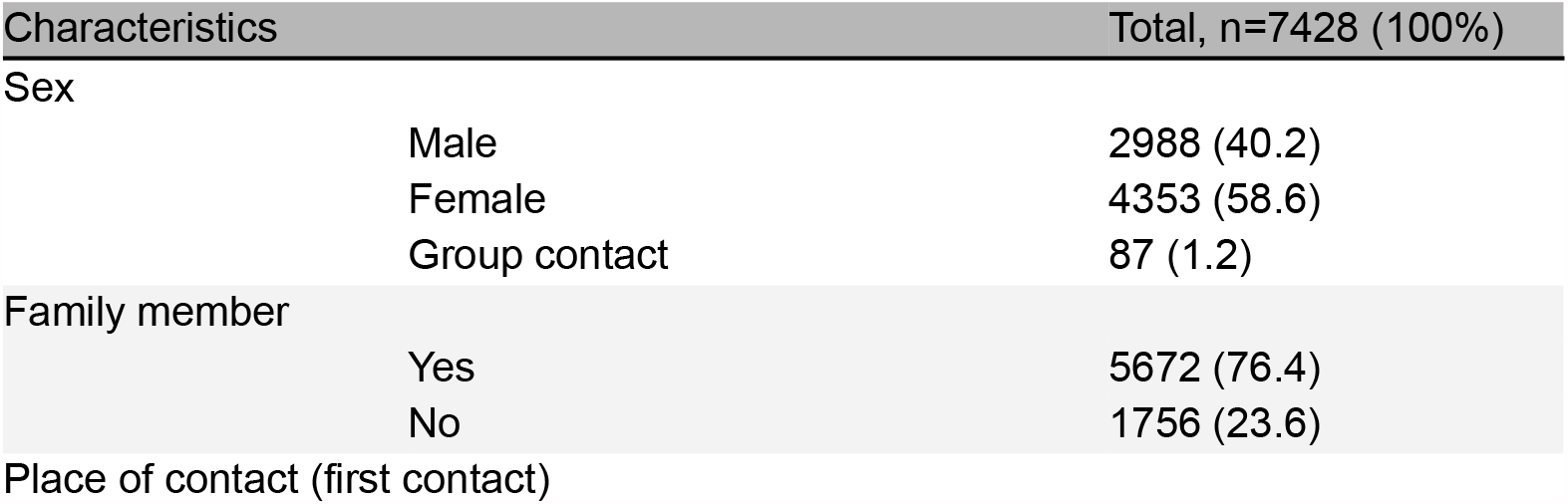
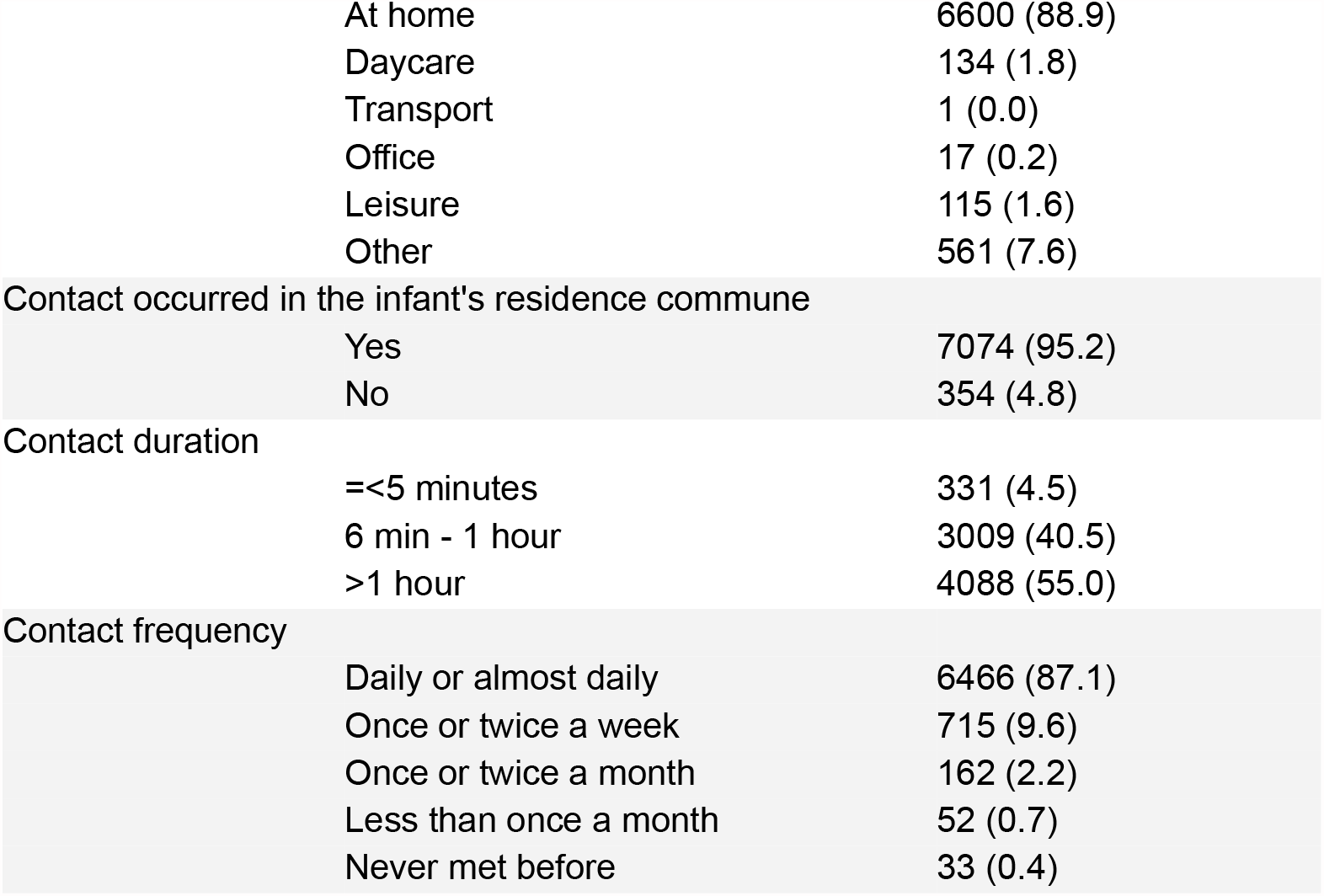
Characteristics of infants’ contacts

| Characteristics |  | Total, n=7428 (100%) |
| --- | --- | --- |
| Sex |  |  |
|  | Male | 2988 (40.2) |
|  | Female | 4353 (58.6) |
|  | Group contact | 87 (1.2) |
| Family member |  |  |
|  | Yes | 5672 (76.4) |
|  | No | 1756 (23.6) |
| Place of contact (first contact) |  |  |

|  |  |
| --- | --- |
| At home | 6600 (88.9) |
| Daycare | 134 (1.8) |
| Transport | 1 (0.0) |
| Office | 17 (0.2) |
| Leisure | 115 (1.6) |
| Other | 561 (7.6) |
| Contact occurred in the infant's residence commune |  |
| Yes | 7074 (95.2) |
| No | 354 (4.8) |
| Contact duration |  |
| =<5 minutes | 331 (4.5) |
| 6 min - 1 hour | 3009 (40.5) |
| >1 hour | 4088 (55.0) |
| Contact frequency |  |
| Daily or almost daily | 6466 (87.1) |
| Once or twice a week | 715 (9.6) |
| Once or twice a month | 162 (2.2) |
| Less than once a month | 52 (0.7) |
| Never met before | 33 (0.4) |

**Figure 1:**
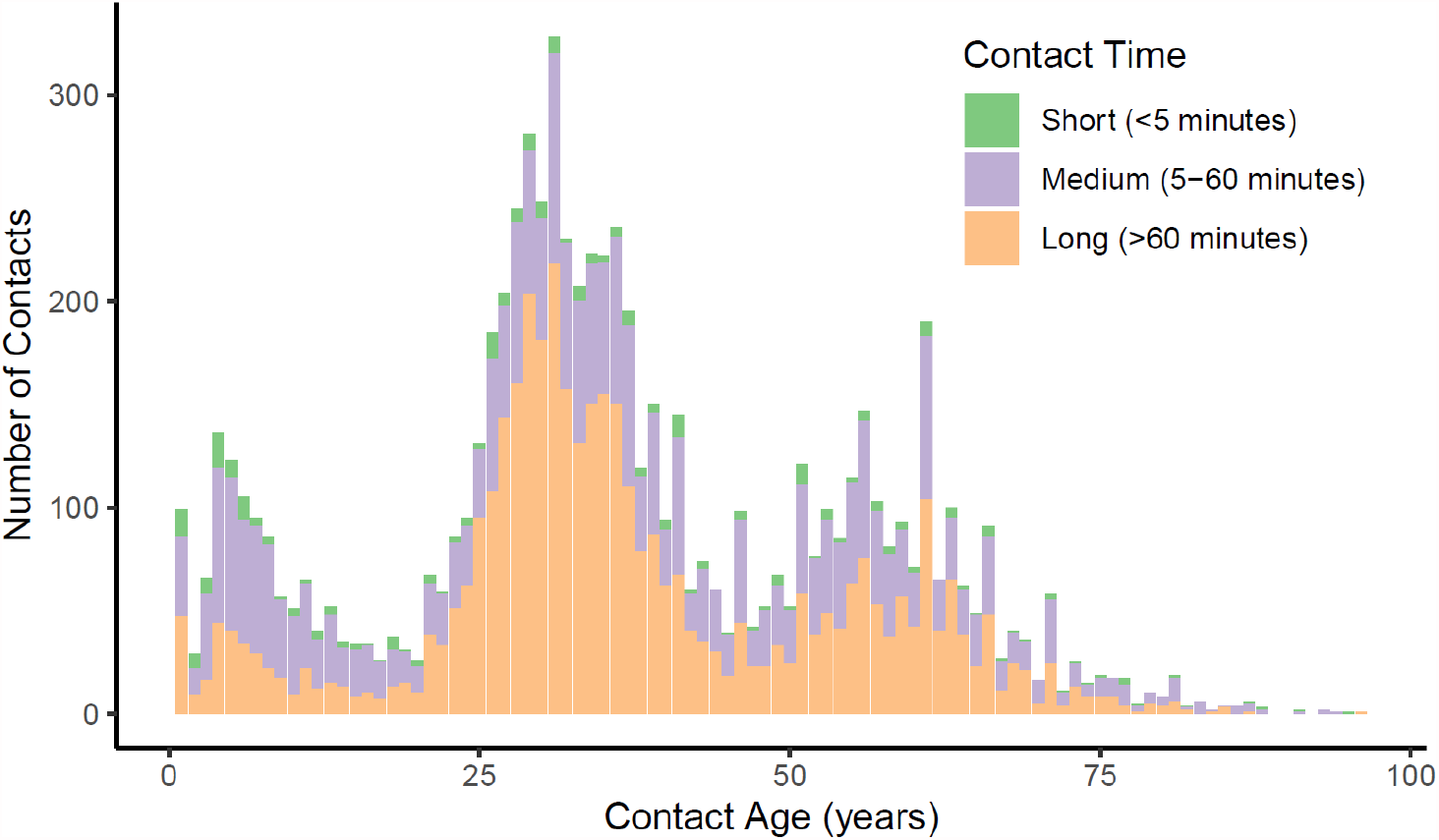
The number of infant contacts. Reported skin to skin contacts are shown disaggregated into annual age groups and stratified by contact duration.

### Factors affecting the number of contacts

The number of skin to skin contacts an infant made on the day before the survey ranged from 1 to 15 and the mean was 4.7 (SD 1.8). Infants who lived in households with more than 4 people were reported to have 25% (19% - 30%) more contacts than infants in smaller households. The infant’s ability to crawl or sit was not found to be associated with their number of contacts; however, infants who had travelled beyond commune borders in the week preceding the survey were reported to have 6% (2% - 10%) more contacts. Infants who were found to carry pneumococci were reported to have slightly more contacts, however, this was not statistically significant (Table 1).

### Factors affecting the location of contacts

About 10% of infants had at least one contact outside of their commune of residence (SITable 3). Infants at least 8 months old and those with caretakers in current employment were 67% (95% CI: 16 - 140) and 67% (95% CI: 19 - 134) more likely to have a contact outside their commune respectively (according to the crude odds ratios). Also, pneumococcal carriers were more likely to have had contact outside the commune; however this was no longer the case if adjusted for day care centre attendance which was strongly associated with contacts outside the commune. Infant to infant contact was rare, only 2% of infants were reported to have contacted another infant in another commune. Of such infant to infant contacts the majority (19/31, 61%) happened in day care.

### Validating contacts as a risk factor for carriage

Modelled carriage prevalence in Nha Trang based on carriage observations from a previous study increased rapidly in infancy and peaked at about 4 years of age. Secondary school age children and particularly adults were rarely carrying pneumococcus; about 10% and less than 5% respectively were infected. Carriage rates observed in 2008 and in 2018 were similar in the age groups observed in both surveys (Figure 2).

**Figure 2:**
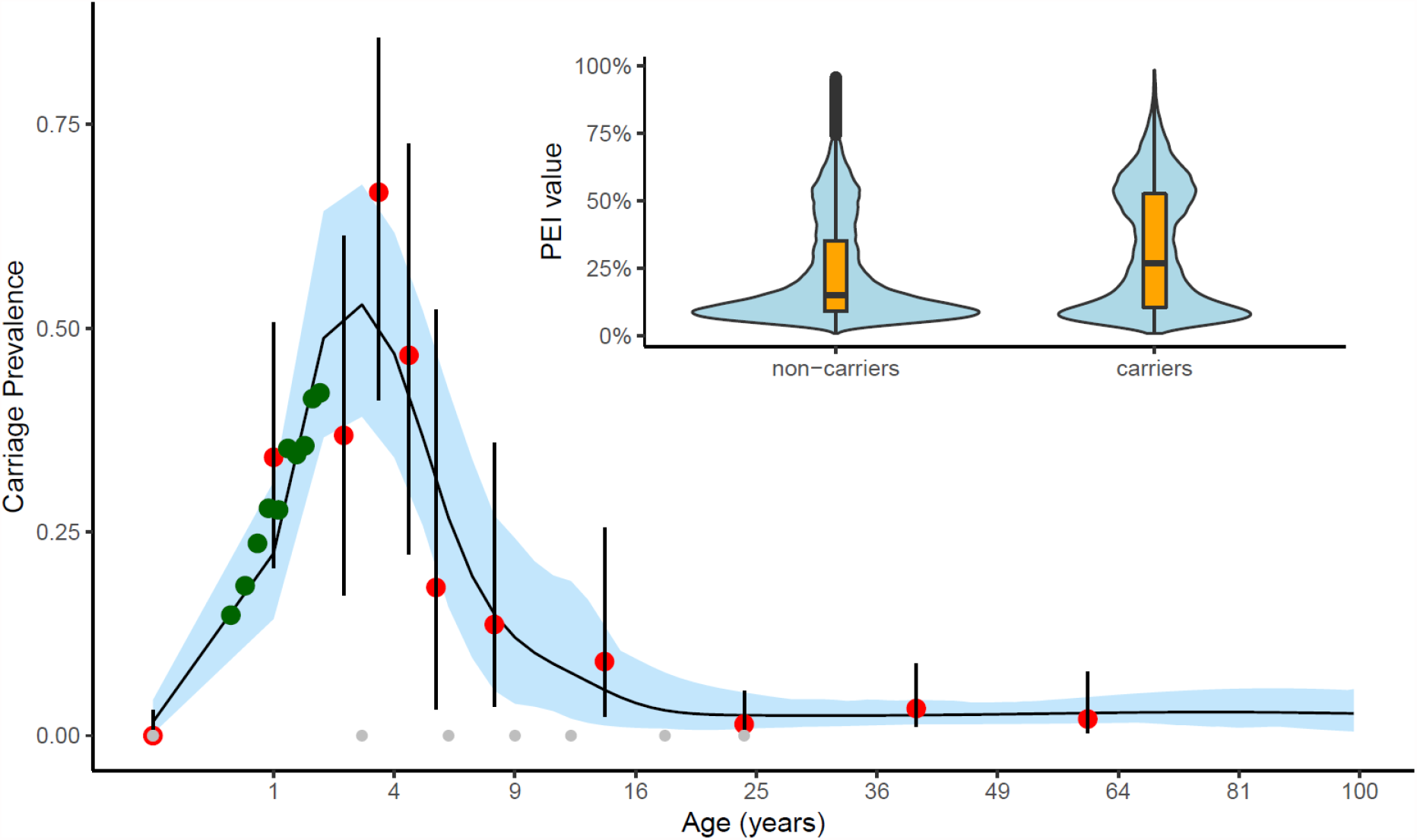
Age-stratified *carriage prevalence in Nha Trang*. Grey dots indicate the positions of knots for the spline and the red dots and their corresponding 95% binomial confidence intervals the carriage prevalence data that the model was fitted to. The black line and the region highlighted in blue represents the model’s median and 95% quantile estimates. The green dots are added for visual comparison and indicate the carriage prevalence observed in the infant survey. To aid visualisation we used quadratic scaling of the x-axis. Inset: the distribution of estimated PEI values in infants without (left) and with (right) pneumococcal carriage.

We estimate that most (61%) infants had less than a 25% probability to have had close contact with a pneumococcal carrier on the surveyed day; however, for 22% of infants the probability of exposure was much higher at 50-70%. The estimated PEI for each infant was associated with substantial uncertainty (SIFigure 5). However, on average pneumococcal carriers had a higher PEI than non carriers; 27% (95% CI: 5.1% - 70%) compared with 15% (95% CI: 4.9% - 61%).

When adjusting for age and locality we find that probability of exposure to pneumococci on the surveyed day and the risk of pneumococcal carriage were highly correlated: for every 10 percentage points increase in PEI the odds of pneumococcal carriage increased by 22% (95% CI: 15 - 29), i.e. the odds of pneumococcal carriage were 7.1 (95% CI: 4.1 - 12.3) times higher in infants who almost certainly had contact with an infected person (PEI ≈ 1) compared to those who had not (SITable 5).

### Age-specific infant exposure to pneumococci

Children between 1 to 4 years of age contributed 38% (22% - 56%) of the total pneumococcal exposure to infants; children 5-9 and adults 21-40 year olds contributed 22% (95% CI: 16-29) and 20% (95% CI: 9.2-32), respectively. We estimate that 2 to 6 year old children are key pneumococcal transmitters to infants, contributing 51% (95% CI: 39-63) of the total exposure, and that infection risk from other infants or older children and adults is likely low in this setting (Figure 3). The majority of potential pneumococcal exposure (80%) originated from household contacts.

**Figure 3:**
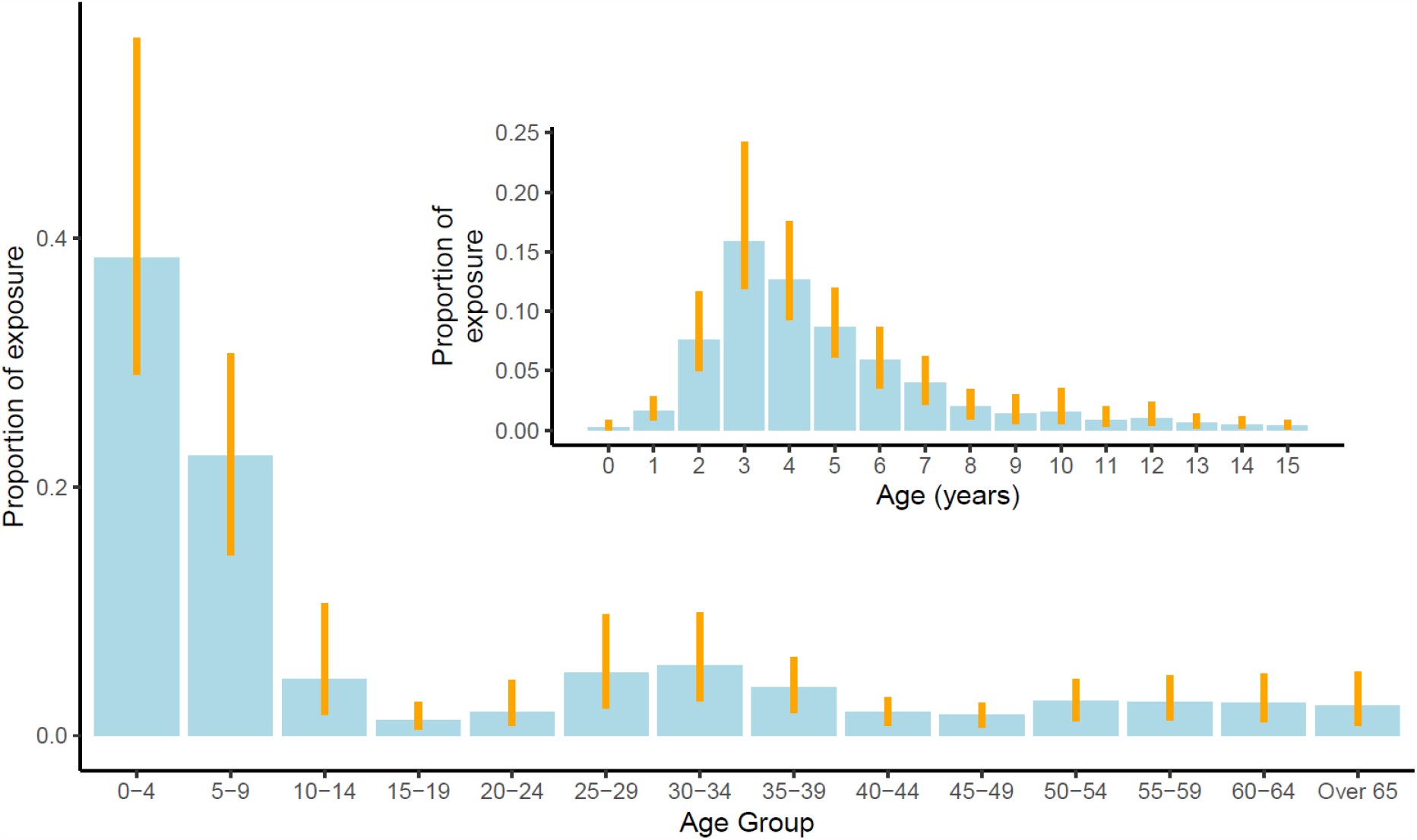
The contribution of different age groups towards the total exposure of infants to pneumococcus. Bars show the relative contribution of an age group to the total pneumococcal exposure (number of close contacts and their propensity to carriage pneumococci) of infants in Nha Trang. Error bars indicate 95% credible intervals. Inset: the contribution of up to 15y olds to total pneumococcal exposure of infants in Nha Trang.

## Discussion

We described in detail the social contacts of infants in Nha Trang, Vietnam. Unlike other age groups, infant contacts are not age assortative and happen largely within the household. We show that indeed close social contacts, as measured in contact surveys on a specific day, and in combination with age specific infection probabilities of such contacts, are highly correlated with the risk of pneumococcal infection of infants in Nha Trang. Thus we estimate that, with 39% (95% CI: 32% to 47%) and 22% (95% CI: 16-29) respectively, 1-4 year olds and 5-9 year olds contribute most to the infection pressure to infants in this setting and that 80% of infections are acquired from household contacts.

We have previously shown that pneumococcal infection risk is correlated with the frequency of physical contacts [5,6]. Here we expand that notion by including the probability that contacts are infected with pneumococci, similar to how age stratified dynamic mathematical models are constructed [3,22,23], and thus provide direct evidence for the value of social contact structure as a proxy for disease transmission routes in mathematical models.

Furthermore, we suggest that the combination of social contacts and age dependent infection probabilities provides a simple and useful method to identify likely transmission routes, without the need for more complex mathematical modelling or the need for longitudinal data collection [24,25].

Similar to their key role in pneumococcal transmission generally [22,26–28], we find that 2-6 year old children are the likely main transmitters of pneumococcal infection to infants. This has important implications for the optimal design of pneumococcal vaccination strategies, particularly those that aim to sustain herd protection while reducing direct protection in infancy [10]: while the exact duration of protection from pneumococcal conjugate vaccines remains relatively uncertain [29], booster dose schedules may induce longer lasting protection and hence may be preferred in settings with high carriage prevalence in older children and hence a potentially high contribution to the pneumococcal infection pressure from that age group [30].

We find that 76% of contacts and 80% of potential pneumococcal exposures to the infants in this study were with members of their households. In contrast, in the UK the household contacts had a less dominant role and 11 weeks to 12 months old infants were reported to have less than 50% of their contacts at home [12]. This illustrates the importance of context in the evaluation of transmission routes for infants. For example, while in the UK many children will attend nursery from 6 to 8 months of age, in Vietnam publicly funded nurseries will only accept children older than 12 months. Thus infant infection routes may be substantially different in settings where childcare responsibilities are shared with older generations or siblings.

Our study suggests that pneumococcal exposure risk to infants stems mostly from household contacts and particularly siblings. In combination with the commune organised schooling system in Vietnam this suggests that the commune is a major spatial entity in determining pneumococcal infection in infants in Nha Trang. The main mode of transport for families in this community was travel by motorcycle, with the vast majority of families owning and using one for travel. The infants were mostly able to sit but fewer were able to crawl or walk (55% and 12%, respectively), and so for individuals who had not achieved these milestones, contact would likely be initiated by parents or siblings.

A limitation of this study is that there is large uncertainty around the carriage rates in teenagers. However, direct contact with teenagers was rare for our study population and hence the associated uncertainty in infection probability does not impact on the overall results much. In settings where teenagers share childcare responsibilities and where high pneumococcal infection rates extend well into teenage years [31–33], such contacts may contribute a larger part to the infection pressure on infants. A further limitation is that contacts were only surveyed on a single day and that carriage status was only assessed about a week after that. The strong correlation between contact behaviour and pneumococcal carriage in our study suggests that, indeed, the contact behaviour on a given day is somewhat representative for the general contact behaviour of that infant. This is an important validation for the adequacy of social contact surveys for informing mathematical transmission models advising public health decision making. Our survey period did not include any major holidays such as the 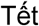 Nguyên Ðán, or Lunar New Year. Such events may well change contact patterns and accelerate the spread of pneumococci (in infants).

In summary, we provide direct evidence that for pneumococci, and thus likely other respiratory pathogens, social contact surveys indeed add a crucial part in our understanding of transmission pathways. We find that 2-6 year old children are the most likely source of pneumococcal infection in infants in Nha Trang. Similar studies in other settings can help evaluate local pneumococcal transmission routes and hence provide crucial evidence for the discussion on optimal dosing schedules for pneumococcal conjugate vaccines.

## Data Availability

Relevant data used in the manuscript can be found in this repository:
https://github.com/GeorgeYQian/Nha-Trang-Contact-Study-Data-and-Code

## Acknowledgements

We would like to thank the staff of Khanh Hoa Health service, and Pasteur Institute in Nha Trang for their support in conducting the contact survey, and the pneumococcal carriage survey. We thank Prof.Kim Mulholland, Prof.Catherine Satzke and team from MCRI for supporting the pneumococcal microarray testing of the samples.

## Author contributions

Conceived field studies: LMY, SF; conceived analysis: AJvH, SF; data analyses: GQ, MT, SC; first draft: GQ, MT, SF; all authors have read and approved the manuscript.

## Supplementary material

**SIFile 1:** Background questionaire

https://drive.google.com/file/d/1YzRA2O_U91mSz5hu6A8gF31LFAxWiID7/view?usp=sharing

**SIFile 2:** Contact questionaire

https://drive.google.com/file/d/1C4xiRwszT7otMReUnC0tG_V-YSqT1E27/view?usp=sharing

**SIFile 3:** Other supporting material:

https://docs.google.com/document/d/1Qj23xe0rN4mUXlFB5Rkp_wOMq4H2ERKZFOdfSBSV8TM/edit?usp=sharing

