## Supplementary Materials for "Pneumococcal exposure routes for infants, a nested cross-sectional survey in Nha Trang, Vietnam"

*Logistic Regression Equation*

The equation for the logistic regression model predicting carriage in infants (see the 'Statistical Analyses' part of the Methods section in the main text) is:

$$logit(p_i) = b_0 + C_i + b_1 E_i + b_2 A_i \quad (1)$$

where:

- $p_i$ is the probability that the *i*th infant in the study is a pneumococcal carrier
- $b_0$ is the model intercept under a single-level regression model (fixed effects only)
- $C_i$ is the random effect on the intercept due to the influence of the commune to which the *i*th individual belongs
- $b_1$ is the coefficient corresponding to the effect of PEI
- $E_i$ is the value of PEI,for the *i*th individual
- $b_2$ is the coefficient corresponding to the effect of the infant's age

- $A_i$ is the age of the *i*th individual in months

### *Model Comparison*

We compared the model in Equation 2 with other logistic regression models with different covariants:

- Carriage ~ Number of Contacts (traditionally the covariate used for predicting carriage)
- Carriage ~ PEI (to justify the inclusion of the other covariates)
- Carriage ~ PEI + Infants' Age
- Carriage ~ PEI + Commune
- Carriage ~ [(PEI + Infants' Age) with commune as groups]|longer duration contacts only (including only long-duration contacts)

These models were compared using the Deviance Information Criterion (DIC) values, a generalisation of the Akaike Information Criterion that is suitable for comparison of Bayesian models, provided that the covariates approximately follow a Gaussian distribution; we found that this was indeed the case for the covariates in our model.
The results are shown in Table 5. The model with PEI as the lone covariate appears to outperform the corresponding model with the number of contacts alone, based on DIC values. The set of models with PEI together with covariates such as mobility and motorcycle share similar DIC values and appear to be the best-performing models; this set includes the proposed model (Equation 2). Including only longer-duration contacts does not appear to reduce the DIC value.
.

**Table S3**: Effect of each infant's characteristic on having contact outside of the commune of residence, estimated using logistic regression model.

| Characteristics | N | % Infants with contacts in other communes | Odds ratio | Adjusted odds ratio* | Adjusted odds ratio** |
|---|---|---|---|---|---|
| Total | 1583 | 154 (9.7) | | | |
| Demographics | | | | | |
| Sex | | | | | |
| Male | 871 | 85 (9.8) | reference | reference | |
| Female | 712 | 69 (9.7) | 0.99 (0.71-1.39) | 0.96 (0.66-1.41) | |
| Age (months) | | | | | |
| <8 months | 616 | 44 (7.1) | reference | | reference |
| 8-13 months | 967 | 110 (11.4) | 1.67 (1.16-2.40) | | 1.41 (1.05-1.90)*** |
| Family | | | | | |
| Siblings in the household | | | | | |
| No sibling | 666 | 66 (9.9) | reference | reference | |
| One or more | 917 | 88 (9.6) | 0.97 (0.69-1.35) | 0.96 (0.74-1.23) | |

| | | | | | |
|---|---|---|---|---|---|
| siblings | | | | | |
| Number of people in the household | | | | | |
| 2-4 | 611 | 62 (10.2) | reference | reference | |
| >4 | 972 | 92 (9.5) | 0.93 (0.66-1.3) | 0.91 (0.62-1.36) | |
| Caretaker currently in a paid employment | | | | | |
| No | 1056 | 86 (8.1) | reference | reference | reference |
| Yes | 527 | 68 (12.9) | 1.67 (1.19-2.34) | 1.60 (1.13-2.27) | 1.08 (0.71-1.65) |
| Highest level of education in the household | | | | | |
| Secondary school or lower | 268 | 27 (10.1) | reference | reference | |
| Degree | 1315 | 127 (9.7) | 0.95 (0.62-1.48) | 0.94 (0.65-1.36) | |
| Infant's activity | | | | | |
| Sit | | | | | |
| Yes | 1334 | 139 (10.4) | 1.81 (1.05-3.15) | 1.36 (0.70-2.62) | |
| No | 249 | 15 (6.0) | reference | reference | |
| Crawl | | | | | |
| Yes | 873 | 95 (10.9) | 1.35 (0.96-1.9) | 1.08 (0.73-1.59) | |
| No | 710 | 59 (8.3) | reference | reference | |
| Walk | | | | | |
| Yes | 186 | 28 (9.0) | 1.79 (1.15-2.78) | 1.51 (0.95-2.39) | |
| No | 1397 | 126 (15.1) | reference | reference | |
| Mobility | | | | | |
| Bicycle | | | | | |
| Yes | 31 | 1 (3.2) | 0.30 (0.04-2.25) | 0.30 (0.04-2.32) | |
| No | 1552 | 153 (9.9) | reference | reference | |
| Motorbike | | | | | |
| Yes | 1549 | 152 (9.8) | 1.74 (0.41-7.34) | 1.73 (0.30-9.9) | |
| No | 34 | 2 (5.9) | reference | reference | |
| Car | | | | | |
| Yes | 99 | 12 (12.1) | 1.3 (0.7-2.44) | 1.34 (0.78-2.31) | |
| No | 1484 | 142 (9.6) | reference | reference | |
| Walk | | | | | |
| Yes | 108 | 10 (9.3) | 0.94 (0.48-1.85) | 0.97 (0.50-1.87) | |
| No | 1475 | 144 (9.8) | reference | reference | |
| Public transportation | | | | | |
| Yes | 40 | 5 (12.5) | 1.34 (0.52-3.46) | 1.44 (0.68-3.03) | |
| No | 1543 | 149 (9.7) | reference | reference | |
| Number of times caretaker left the commune in the last 7 days | | | | | |
| 0-2 | 872 | 44 (5.1) | reference | reference | reference |
| 3 or more | 711 | 110 (15.5) | 3.44 (2.39-4.96) | 3.36 (2.02-5.58) | 3.01 (1.78-5.10) |
| Number of times infant left the commune in the last 7 days | | | | | |
| 0 | 788 | 9 (1.1) | reference | reference | reference |
| 1 or more | 795 | 145 (18.2) | 19.31 (9.77-38.16) | 18.9 (7.99-44.72) | 18.3 (7.87-42.47) |

| | | | | | |
|---|---|---|---|---|---|
| Day-care | | | | | |
| Day-care attendance | | | | | |
| Yes | 1497 | 121 (8.1) | 7.08 (4.41-11.36) | 6.56 (3.92-10.98) | |
| No | 86 | 33 (38.4) | reference | reference | |
| Pneumococcus carriage (n=1582) | | | | | |
| Pneumococcus | | | | | |
| Yes | 353 | 48 (13.6) | 1.67 (1.16-2.40) | 1.58 (1.13-2.21) | 1.25 (0.92-1.71) |
| No | 1229 | 106 (8.6) | reference | reference | reference |

*Odds ratios adjusted by age group, considering clustering in each commune
**Odds ratios adjusted by age group & day-care attendance, considering clustering in each commune
***Odds ratio adjusted by day-care attendance, considering clustering in each commune

**Table S4:** A comparison of different logistic regression models to predict carriage, using DIC values

| Covariates | DIC value |
|---|---|
| Number of Contacts | 1682 |
| PEI | 1656 |
| PEI + Infants' Age | 1618 |
| PEI+Infants' Age, with Commune as a random effect | 1618 |
| PEI + Infants' Age - Longer Duration Contacts Only, with Commune as a random effect | 1619 |

**Table S5:** Values of the coefficients from the logistic regression (Equation 2) are given here.

| Parameter | Mean Value | 2.5% | 97.5% |
|---|---|---|---|
| Fixed-effects model Intercept ($\beta_0$) | -3.33 | -4.82 | -2.05 |
| Effect of PEI ($\beta_1$ ) | 1.96 | 1.40 | 2.51 |
| Commune ID: 21 ($C_1$) | 0.13 | -0.39 | 0.64 |
| Commune ID: 17 ($C_2$) | 0.21 | -0.32 | 0.71 |
| Commune ID: 16 ($C_3$) | 0.06 | -0.46 | 0.57 |

| | | | |
|---|---|---|---|
| Commune ID: 23 ($C_4$) | 0.01 | -0.52 | 0.53 |
| Commune ID: 18 ($C_5$) | -0.11 | -0.68 | 0.43 |
| Commune ID: 14 ($C_6$) | 0.18 | -0.32 | 0.70 |
| Commune ID: 27 ($C_7$) | -0.12 | -0.66 | 0.41 |
| Commune ID: 10 ($C_8$) | -0.16 | -0.71 | 0.37 |
| Commune ID: 7 ($C_9$) | 0.05 | -0.50 | 0.58 |
| Commune ID: 3 ($C_{10}$) | -0.43 | -1.09 | 0.14 |
| Commune ID: 25 ($C_{11}$) | -0.40 | -1.03 | 0.17 |
| Commune ID: 20 ($C_{12}$) | 0.39 | -0.13 | 0.90 |
| Commune ID: 13 ($C_{13}$) | 0.11 | -0.43 | 0.61 |
| Commune ID: 11 ($C_{14}$) | -0.02 | -0.58 | 0.52 |
| Commune ID: 5 ($C_{15}$) | -0.16 | -0.73 | 0.35 |
| Commune ID: 15 ($C_{16}$) | 0.55 | 0.05 | 1.05 |
| Commune ID: 24 ($C_{17}$) | 0.32 | -0.20 | 0.84 |
| Commune ID: 12 ($C_{18}$) | -0.66 | -1.34 | -0.07 |
| Commune ID: 1 ($C_{19}$) | -0.15 | -0.71 | 0.38 |
| Commune ID: 4 ($C_{20}$) | 0.01 | -0.51 | 0.50 |
| Commune ID: 19 ($C_{21}$) | -0.37 | -0.96 | 0.17 |
| Commune: ID 22 ($C_{22}$) | 0.09 | -0.45 | 0.60 |
| Commune ID: 2 ($C_{23}$) | -0.17 | -0.73 | 0.36 |
| Commune ID: 9 ($C_{24}$) | 0.21 | -0.33 | 0.74 |
| Commune ID: 8 ($C_{25}$) | 0.04 | -0.50 | 0.55 |
| Commune ID: 6 ($C_{26}$) | -0.17 | -0.74 | 0.36 |

| | | | |
|---|---|---|---|
| Commune ID: 26($C_{27}$) | -0.12 | -0.72 | 0.45 |
| Infants' Age ($b_2$) | 0.04 | 0.01 | 0.08 |

**Table S6:** The VIF values quantifying multicollinearity among the covariates of the model are shown here. The values are well below 10 (which, as a 'rule of thumb', is when multicollinearity becomes influential).

| Covariate | VIF |
|---|---|
| Mean PEI value | 1.05 |
| Infant Age | 1.03 |
| Commune | 1.08 |

**Figure S4:** The Directed Acyclic Graph of the factors involved in the spread of pneumococcus to infants in Nha Trang is shown here

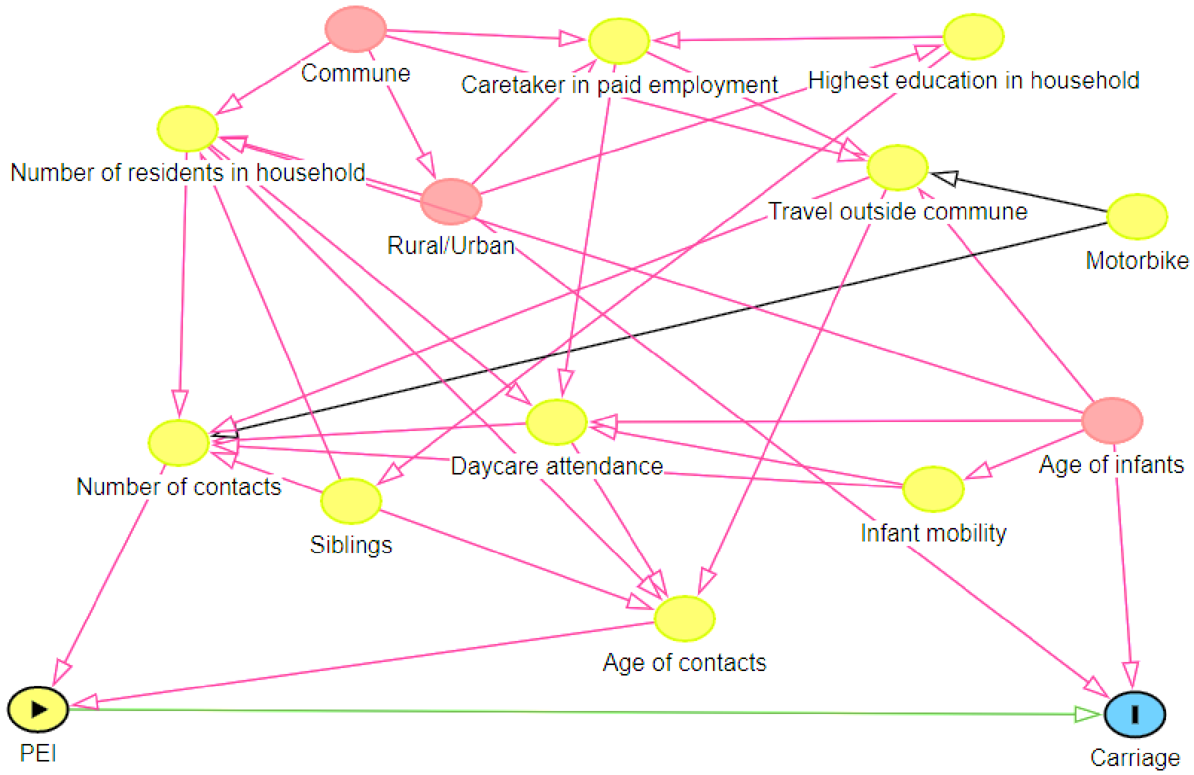

**Figure S5:** Distribution of 10000 MCMC samples of PEI for a randomly selected individual (Individual A), shown through a violin plot and boxplot nested within.

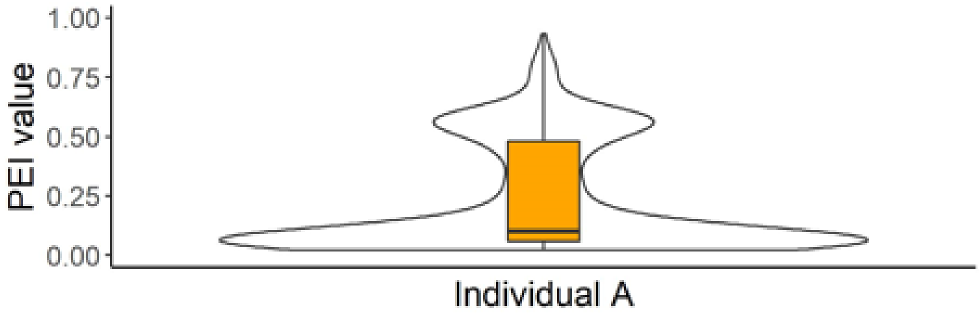
